# SARS-CoV-2 seroprevalence in a rural and urban community household cohort in South Africa, after the third wave, April-November 2021

**DOI:** 10.1101/2022.02.10.22270772

**Authors:** Jackie Kleynhans, Stefano Tempia, Nicole Wolter, Anne von Gottberg, Jinal N. Bhiman, Amelia Buys, Jocelyn Moyes, Meredith L. McMorrow, Kathleen Kahn, F. Xavier Gómez-Olivé, Stephen Tollman, Neil A. Martinson, Floidy Wafawanaka, Limakatso Lebina, Jacques du Toit, Waasila Jassat, Mzimasi Neti, Marieke Brauer, Cheryl Cohen, the PHIRST-C Group

**Author notes:** **Address for correspondence** Jackie Kleynhans, Centre for Respiratory Diseases and Meningitis, National Institute for Communicable Diseases of the National Health Laboratory Service, 1 Modderfontein Road, Sandringham, 2192, Johannesburg, South Africa. Additional members of the PHIRST-C group who contributed to this manuscript are listed at the end of this article.

## Abstract

By November 2021, after the third SARS-CoV-2 wave in South Africa, seroprevalence was 60% (95%CrI 56%-64%) in a rural and 70% (95%CrI 56%-64%) in an urban community; highest in individuals aged 13-18 years. High seroprevalence prior to Omicron emergence may have contributed to reduced severity observed in the 4^th^ wave.

**Article Summary Line:** In South Africa, after a third wave of SARS-CoV-2 infections, seroprevalence was 60% in a rural and 70% in an urban community, with case-to-infection, - hospitalization and -fatality ratios similar to the second wave.

---

South Africa so far experienced four waves of SARS-CoV-2 infection, the fourth dominated by the Omicron variant of concern (1). Data on the proportion of the population with serologic evidence of previous infection at the time of Omicron emergence are important to contextualise the observed rapid increases and subsequent quick decline in case numbers, (1) as well as the lower severity compared to previous variants (2).

We previously described the seroprevalence of SARS-CoV-2 in the PHIRST-C cohort in a rural and an urban community at five time points, from July 2020 to March 2021 (3). Using the same methods (Appendix), we report seroprevalence at four additional time points through November 27, 2021, spanning the third Delta-dominated wave (Appendix Figure 1), ending the week Omicron was identified (4). We tested sera using the Roche Elecsys® Anti-SARS-CoV-2 assay (Roche Diagnostics, Rotkreuz, Switzerland); cut-off index (COI)≥1.0 considered indication of prior infection. The immunoassay detects nucleocapsid (N) antibodies, thus not detecting post-vaccination antibody responses. We obtained seroprevalence 95% credible intervals (CrIs) by using Bayesian inference with 10,000 posterior draws (5). We estimated the age-and sex-adjusted number of infections, and age-adjusted diagnosed cases, hospitalizations, deaths, case-to-infection ratio (CIR), case-to-hospitalization ratio (HIR) and in-hospital and excess death case-to-fatality ratio (FIR) as described previously (3) and in Appendix. Third wave infections were defined as participants with a paired blood draw (BD) 5 (collected March 22 – April 11, 2021) and BD9 (collected November 15 – 27, 2021) sample, seronegative at BD5 and seropositive at BD9, or seropositive at BD5 but had a ≥2-fold higher COI in BD9 (as possible re-infections after BD5 occurred, n=38, detailed in Appendix). Vaccination status was obtained through reviewing vaccine cards kept at home. The study was approved by the University of the Witwatersrand Human Research Ethics Committee (Reference 150808) and the US Centers for Disease Control and Prevention relied on local clearance (IRB #6840).

Overall, pre-third wave (BD5) SARS-CoV-2 seroprevalence adjusted for assay sensitivity and specificity was 26% (95% CrI 22%-29%) in the rural and 41% (95% CrI 37%-45%) in the urban community. After the third wave (BD9), overall seroprevalence increased to 60% (95% CrI 56%-64%) in the rural community and 70% (95% CrI 66%-74%) in the urban community (Figure, Appendix Table 1). In both communities the largest increase in seroprevalence was seen in children aged 13-18 years, who also had the highest seroprevalence of all ages after the third wave: 80% (95% CrI 70%-88%) in the rural community and 83% (95% CrI 73%-90%) in the urban community, a 49% and 19% increase, respectively.

**Figure.**
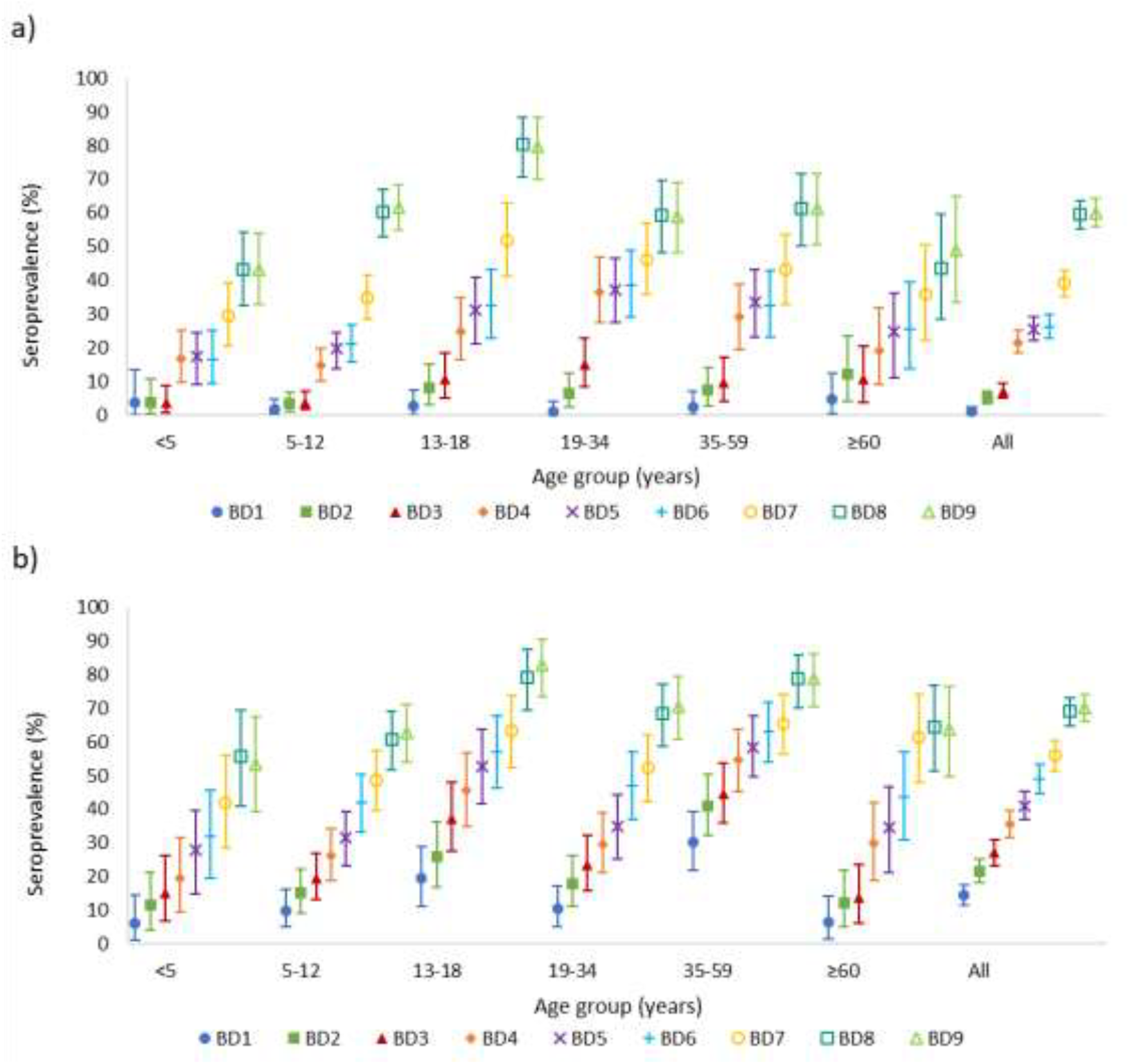
SARS-CoV-2 seroprevalence at each blood collection by age group in the a) rural and b) urban community March 2020 - November 2021, South Africa. Baseline (BD1) July 20 – September 17, 2020; second draw (BD2) September 21 – October 10, 2020; third draw (BD3) November 23 – December 12, 2020; fourth draw (BD4) January, 25 – February 20, 2021; fifth draw (BD5) March 22 – April 11, 2021; sixth draw (BD6) May 20 – June 9, 2021; seventh draw (BD7) July 19 – August 5, 2021; eighth draw (BD8) September 13 – 25, 2021; ninth draw (BD9) November 15 – 27, 2021. Vertical capped lines represent 95% credible interval. Seroprevalence estimates adjusted for sensitivity and specificity of assay.

During the third wave of infections, the incidence at the rural site was 39% (95%CrI 24%-55%), resulting in a CIR of 3% (95%CI 2%-5%). There was a 0.5% (95%CI 0.3%-0.7%) HIR and an in-hospital FIR of 0.1% (95%CI 0.1%-0.2%) and an excess deaths FIR of 0.5% (95%CI 0.4%-0.8%, Figure and Appendix Figure 2).

In the urban community the incidence during the third wave was 40% (95%CrI 26%-54%). There was a 5% (95%CI 4%-8%) CIR and 2% (95%CI 2%-4%) HIR. The in-hospital FIR was 0.4% (95%CI 0.3%-0.6%) and excess deaths FIR 0.6% (95%CI 0.4-0.9%, Figure and Appendix Figure 2).

At the time of BD9, for those individuals in the rural community with a known vaccination status, 29% (25/85) of individuals aged 35-59 years, and 38% (16/42) aged ≥60 years were fully vaccinated against SARS-CoV-2 (one dose of Johnson & Johnson or two doses of Pfizer-BioNTech). In the urban community, 42% (47/111) aged 35-59 years and 58% (31/53) aged ≥60 years were fully vaccinated (Appendix Table 2).

Taken together, by the end of November 2021, just prior to the emergence of Omicron, the combined proportion of individuals with either serologic evidence of previous infection (at any draw) and/or fully vaccinated was 62% (389/631) at the rural and 72% (411/568) at the urban community (Appendix Table 3).

After the third wave of infections in South Africa, we observed a ≥60% overall seroprevalence due to SARS-CoV-2 infection, ranging from 43% in rural community children aged <5 years to 83% in urban community children aged 13-18 years (Figure). CIF, HIR and FIRs were similar between the second and third waves. Similar to our data, a study from Gauteng Province found seroprevalence of 56%-80% due to natural infection prior to the emergence of Omicron (6). The high seroprevalence prior to Omicron emergence may have contributed to reduced severity observed in the 4^th^ wave (2). Future studies examining SARS-CoV-2 attack rates using paired serology will be important to help understand how immunity affects disease severity in the population during the fourth Omicron wave.

## Data Availability

The investigators welcome enquiries about possible collaborations and requests for access to the dataset. Data will be shared after approval of a proposal and with a signed data access agreement. Investigators interested in more details about this study, or in accessing these resources, should contact the corresponding author.

## Additional members of the PHIRST-C group who contributed to this manuscript

Kgaugelo Patricia Kgasago, Linda de Gouveia, Maimuna Carrim, Mignon du Plessis, Retshidisitswe Kotane, Tumelo Moloantoa

## Acknowledgements

All individuals participating in the study, field teams for their hard work and dedication to the study, the laboratory teams, the PHIRST-C scientific and safety committee, the national SARS-CoV-2 NICD surveillance team, NICD IT.

## Disclaimer

The findings and conclusions in this report are those of the authors and do not necessarily represent the official position of the CDC.

## Funding

This work was supported by the National Institute for Communicable Diseases of the National Health Laboratory Service and the US Centers for Disease Control and Prevention (cooperative agreement number 6U01IP001048-04-02 awarded to C Cohen). The funders had no role in study design, data collection and analysis, decision to publish, or preparation of the manuscript.

## Potential conflicts of interest

Cheryl Cohen reports receiving grant funds from US-Centers for Disease Control and Prevention, Wellcome Trust and South African Medical Research Council. Nicole Wolter and Anne von Gottberg report receiving grant funds from Sanofi and Gates Foundation. All other authors have no competing/conflict of interest.

## Biographical Sketch

Jackie Kleynhans is an epidemiologist in Centre for Respiratory Diseases and Meningitis, National Institute for Communicable Diseases. She holds a Master’s degree in Microbiology and Masters of Public Health from the University of Pretoria, and is an alumnus of the South African Field Epidemiology Training Programme. Her interests include the epidemiology of respiratory diseases like influenza and COVID-19, vaccine impact studies and modelling of infectious disease transmission dynamics.

## Address for correspondence

Jackie Kleynhans, Centre for Respiratory Diseases and Meningitis, National Institute for Communicable Diseases of the National Health Laboratory Service, 1 Modderfontein Road, Sandringham, 2192, Johannesburg, South Africa;

## Notes

### Author Declarations

The study was approved by the University of the Witwatersrand Human Research Ethics Committee (Reference 150808) and the US Centers for Disease Control and Prevention relied on local clearance (IRB #6840).

