## Appendix for "SARS-CoV-2 seroprevalence in a rural and urban community household cohort in South Africa, after the third wave, April-November 2021"

### Supplementary Methods

#### Study Population

PHIRST-C (A Prospective Household study of SARS-CoV-2, Influenza, and Respiratory Syncytial virus community burden, Transmission dynamics and viral interaction in South Africa) is a continuation from PHIRST (A Prospective Household observational cohort study of Influenza, Respiratory Syncytial virus and other respiratory pathogens community burden and Transmission dynamics in South Africa) (1). PHIRST-C is conducted in two communities with established influenza-like illness and pneumonia surveillance sites.

South Africa consists of nine provinces, which are divided into 52 districts, which form the second level of administration. Districts are further divided into municipalities. The rural site is located in the Bushbuckridge Municipality, Ehlanzeni District, Mpumalanga Province and forms part of a health and sociodemographic surveillance site (HDSS) at the Medical Research Council (MRC)/University of Witwatersrand Rural Public Health and Health Transitions Research Unit, Agincourt. During PHIRST, two of the 29 villages within the HDSS were selected according to convenience (proximity and burden of other studies within the site) each year between 2016–2018. A random selection of ≈50 households with a household size of 3 or more were approached for enrolment. The urban site is located in the Jouberton Township, Matlosana Municipality, Dr Kenneth Kaunda District, North West Province. A list of 450 global positioning system (GPS) coordinates were generated using Google Earth. Study staff approached the nearest house within 30 m of the coordinate point for enrolment. Approximately 50 households were enrolled each year during 2016–2018. At both sites, households were eligible if they consisted of 3 or more household members (sharing at least four meals a week). All household members were approached for inclusion in the study, and households were eligible for inclusion if >80% of members consented to be enrolled.

For PHIRST-C, all households who participated in PHIRST from the urban site, and households from the 2017 and 2018 cohort in the rural site were approached for enrolment. To supplement the sample size, additional households at each site were approached using the same methods as for PHIRST. Villages in the rural site were restricted to those four used in 2017 and 2018 of PHIRST. Informed consent was obtained from participating adults or a parent/guardian for children <18 years of age. In addition to parent/guardian consent, assent was also obtained from children aged 7–17 years.

We collected baseline data and blood (blood draw [BD] 1) at enrollment (July 20–September 17, 2020) and every 2 months thereafter: BD2, September 21–October 10; BD3, November 23–December 12, 2020; BD4, January 25–February 20, 2021; BD5, March 22–April 11, 2021; BD6, May 20 – June 9, 2021; BD7, July 19 – August 5, 2021; BD8, September 13 – 25, 2021; BD9, November 15 – 27, 2021.

The Bushbuckridge Municipality within the Ehlanzeni District is considered as predominantly rural, with main industries being agriculture and tourism (2). The Ehlanzeni District has a population of 1,828,738, with sixty percent of the district's population being >18 years of age. The City of Matlosana Municipality is considered to be 88.2% urban, and is located in the Dr Kenneth Kaunda District, which has a population of 797,715 (3). Sixty-five percent of the district population is aged >18 years.

#### Calculation of Case-to-infection Ratio (CIR), Hospitalization-to-infection Ratio (HIR), and Fatality-to-infection Ratio (FIR) by Wave of Infection

We calculated the age- and sex-adjusted total number of infections, age-adjusted diagnosed cases, hospitalizations, deaths, CIR (number of infections compared to diagnosed cases), HIR and in-hospital and excess death FIR as described in below equations. We defined the third wave as March 22, 2021 (week 12) to November 14, 2021 (week 45), starting with BD5, and ending the week before BD9 started. Due to differences between the sex ratios of our cohort and the district population, the infection estimates were also adjusted for sex. Data sources used in these calculations are described in the next section. Age- and sex- standardized estimates for the selected endpoints for each wave were obtained as follows:

$$Infections = \frac{\sum_i (s_i \times SAp_i)}{\sum_i (SAp_i)} \times 100{,}000$$

Where $s_i$ is the seroprevalence in the cohort in the respective community for age and sex group $i$ and $SAp_i$ is the South African population for age and sex group $i$. Calculated for wave 3 as the number of individuals seronegative at BD5 and seropositive at BD9. Furthermore, we also included possible re-infections where the individual was already seropositive at BD5, but had a ≥2-fold higher cut-off index (COI) in BD9 compared to BD5. We therefore included infections and re-infections that occurred during the third wave. Estimates only included participants with a blood draw 5 and 9 pair, and adjusted for sensitivity and specificity of test (4).

The ≥2-fold increase in COI cutoff to define possible re-infections was not externally validated, but based on PCR-confirmed infections from twice-weekly nasal swab collections between March 22, 2021 (BD5) to July 19, 2021 (BD7) in the same cohort (5). Increases in COI from BD5 to BD7 ranged from 1 to 93 fold. Of the 30 individuals who had a rRT-PCR-confirmed infection between BD5 and BD7, 16 (53%, sensitivity) had a ≥2-fold increase from BD5 to BD7, and of the 312 individuals that did not have a rRT-PCR infection, 301 (96%, specificity) did not have a ≥2-fold increase. We were therefore more likely to have underestimated re-infections.

$$Diagnosed\ cases = \frac{\sum_i \left( (c_i \div p_{i(d)}) \times p_{i(SA)} \right)}{\sum_i \left( p_{i(SA)} \right)} \times 100{,}000$$

Where $c_i$ is the number of diagnosed cases (RT-PCR and antigen-based tests) from the respective district reported to the NMCSS during wave 3 (March 22, to November 14, 2021) in age group $i$, $p_{i(d)}$ is the district population for age group $i$ and $p_{i(SA)}$ is the South African population for age group $i$.

$$Hospitalisations = \frac{\sum_i \left( (h_i \div p_{i(d)}) \times p_{i(SA)} \right)}{\sum_i \left( p_{i(SA)} \right)} \times 100{,}000$$

Where $h_i$ is the number of hospitalizations from the respective district reported to COVID-19 Sentinel Hospital Surveillance (DATCOV) during wave 3 (March 22, to November 14, 2021) (6) in age group $i$, $p_{i(d)}$ is the district population for age group $i$ and $p_{i(SA)}$ is the South African population for age group $i$.

$$In-hospital\ deaths = \frac{\sum_i \left( (d_i \div p_{i(d)}) \times p_{i(SA)} \right)}{\sum_i \left( p_{i(SA)} \right)} \times 100{,}000$$

Where $d_i$ is the number of in-hospital deaths from the respective districts reported to DATCOV during wave 3 (March 22, to November 14, 2021) (6) in age group $i$, $p_{i(d)}$ is the district population for age group $i$ and $p_{i(SA)}$ is the South African population for age group $i$.

$$Excess\ deaths = \frac{(ExD\ x\ 0.85)\ \times\ p_{(SA)}}{p_{(SA)}} \times 100{,}000$$

Where *ExD* is the rate of provincial excess deaths adjusted to the South African population reported by South African Medical Research Council during wave 3 (March 22, to November 14, 2021) (7), and $p_{(SA)}$ is the total South African population. According to estimates only 85% of excess deaths are attributable to COVID-19 (7).

For infections, hospitalizations, in-hospital (minimum) and excess (maximum) deaths, 95% CIs were calculated using the Clopper-Pearson method with a Poisson distribution.

$$Case - to - infection\ ratio\ (CIR) = \frac{Diagnosed\ cases *}{Infections}\ x\ 100$$

$$Hospitalization - to - infection\ ratio\ (HIR) = \frac{Hospitalisations *}{Infections}\ x\ 100$$

$$IFatality\ ratio - to - infection\ (FIR) = \frac{Deaths *}{Infections}\ x\ 100$$

*As calculated above

Confidence intervals for infection ratios were calculated as ratios from the 95% confidence intervals of infection, hospitalization and death rates.

**Data Sources**

Population Denominators (8)

Population numbers for each district, by age, were obtained from the StatsSA 2021 mid-year population estimates.

Notifiable Medical Conditions Surveillance System (NMCSS) (9)

Reverse transcription PCR (RT-PCR) testing in South Africa to detect SARS-CoV-2 RNA started on 28 January 2020. Rapid SARS-CoV-2 antigen testing was implemented in November 2020. All laboratories in the private and public sector performing SARS-CoV-2 tests automatically feed testing data from their lab information systems to the NMCSS, from where daily and weekly reporting on national, provincial and district SARS-CoV-2 infections are performed. No serology tests are used for this reporting. Results from antigen tests may be underestimated in the NMCSS as not all antigen test results are captured on lab information systems which feed into NMCSS. The number of cases reported to the NMCSS in the Ehlanzeni District (rural community) and Dr Kenneth Kaunda District (urban community) was used as an estimate of reported, diagnosed SARS-CoV-2 infections.

##### COVID-19 National Hospital Surveillance (DATCOV) (6)

DATCOV is a national hospital surveillance system to which all hospitals where COVID-19 admissions occurred in the public and private sector in South Africa report. The case definition includes any person admitted with a positive RT-PCR test result for SARS-CoV-2; and may include individuals for whom the main cause of hospitalization was not SARS-CoV-2. In-hospital outcome was available for all patients. The total number of hospitalizations and in-hospital deaths in each district reported to DATCOV between March 22, to November 14, 2021 was used to calculate in the HIR and minimum FIR, respectively.

##### South African Medical Research Council (SAMRC) Report on Weekly Deaths (7)

The Burden of Disease Unit at the SAMRC produces a weekly report on excess deaths in South Africa. Deaths in South Africa are registered on the Department of Home Affair’s National Population Register and includes all citizens with a South African identification number. The number of excess deaths were defined as the number of all-cause deaths in that week minus the number of deaths expected for the week based on 2014–2019 trends. Excess death estimates are adjusted for incomplete reporting, and death estimates in provinces are age-standardized to the national population. Eighty-five percent of the reported excess deaths were used to calculate the maximum HIR as suggested by SAMRC.

##### Limitations

Our study is limited by a small sample size, reducing the power for accurate seroprevalence estimates in small age strata, and inclusion of only 2 geographic sites, including only households with ≥3 people, with high unemployment in selected households compared to the community unemployment rate (10) and therefore may not be representative of other districts and provinces in South Africa. Our definition of re-infections has not been validated elsewhere and is based on a small number of infections that occurred between BD5 and BD7, and did not consider different variants. Based on data from the same cohort, these reinfections occurred in only a small portion (3%) of the cohort (C. Cohen et al., unpub. data, https://doi.org/10.1101/2021.07.20.21260855) and would have had a negligible influence on the infection ratios. CIR, HIR, and FIR formed part of an ecologic analysis, which is inherently prone to biases. Transmission dynamics within our cohort may not be similar to the transmission dynamics within the district. Twenty percent (260/1,277) of persons did not have a BD 5+9 pair, and bias could have been introduced if the seroprevalence were different for those without a BD 5+9 blood pair.

**Appendix Table 1**. Individuals seropositive for SARS-CoV-2 antibodies with adjusted seroprevalence and 95% credible intervals by blood draw, site and age, July 2020 – November 2021, South Africa*

| | | Seropositive/Tested (Seroprevalence*, 95% CrI) | | | | | | | | |
|---|---|---|---|---|---|---|---|---|---|---|
| Site | Age group, y | BD1 | BD2 | BD3 | BD4 | BD5 | BD6 | BD7 | BD8 | BD9 |
| Rural | <5 | 0/25 (4, 0-14) | 1/49 (4, 0-14) | 2/78 (4, 1-9) | 14/87 (17, 10-25) | 15/89 (18, 10-26) | 14/88 (17, 10-25) | 26/90 (29, 20-39) | 33/77 (43, 32-54) | 36/84 (43, 33-54) |
| | 5-12 | 2/148 (2, 0-5) | 5/169 (3, 0-5) | 7/195 (4, 2-7) | 29/202 (15, 10-20) | 39/201 (20, 15-25) | 42/202 (21, 16-27) | 70/202 (35, 28-42) | 120/200 (60, 53-67) | 122/198 (62, 55-68) |
| | 13-18 | 1/69 (3, 0-7) | 5/72 (8, 0-7) | 8/80 (11, 5-18) | 20/82 (25, 16-35) | 24/78 (31, 22-41) | 24/75 (32, 23-43) | 40/77 (52, 41-63) | 59/73 (80, 71-88) | 57/71 (80, 70-88) |
| | 19-34 | 0/87 (1, 0-4) | 5/88 (7, 0-4) | 13/91 (15, 8-23) | 33/91 (37, 27-47) | 35/95 (37, 28-47) | 34/89 (39, 29-49) | 40/87 (46, 36-57) | 48/81 (59, 48-70) | 47/80 (59, 48-69) |
| | 35-59 | 1/76 (2, 0-7) | 5/77 (8, 0-7) | 7/80 (10, 4-17) | 23/81 (29, 20-39) | 27/82 (33, 24-44) | 27/84 (33, 23-43) | 35/82 (43, 33-54) | 49/80 (61, 50-72) | 49/80 (61, 50-71) |
| | ≥60 | 1/40 (5, 0-12) | 4/39 (12, 0-12) | 4/44 (11, 4-21) | 8/45 (19, 9-32) | 10/42 (25, 14-39) | 10/41 (26, 14-39) | 14/40 (36, 22-51) | 16/37 (44, 28-60) | 18/37 (49, 33-65) |
| | All | 5/445 (1, 0-2) | 25/494 (5, 0-2) | 41/568 (7, 5-9) | 127/588 (22, 18-25) | 150/587 (26, 22-29) | 151/579 (26, 23-30) | 225/578 (39, 35-43) | 325/548 (59, 55-64) | 329/550 (60, 56-64) |
| Urban | <5 | 2/45 (6, 1-15) | 5/50 (11, 4-21) | 7/50 (15, 7-26) | 8/44 (20, 9-32) | 13/48 (28, 16-41) | 14/45 (32, 19-46) | 19/46 (42, 28-56) | 25/45 (56, 41-70) | 24/45 (53, 39-67) |
| | 5-12 | 11/116 (10, 5-16) | 17/116 (15, 9-22) | 24/125 (20, 13-27) | 32/123 (26, 19-34) | 38/122 (32, 24-40) | 52/125 (42, 33-50) | 60/124 (49, 40-57) | 75/124 (61, 52-69) | 77/123 (63, 54-71) |
| | 13-18 | 14/75 (19, 11-29) | 19/75 (26, 17-36) | 30/81 (37, 28-48) | 35/77 (46, 35-57) | 41/78 (53, 42-64) | 44/77 (57, 46-68) | 47/74 (63, 52-74) | 59/74 (79, 69-88) | 60/72 (83, 73-90) |
| | 19-34 | 10/102 (10, 5-17) | 17/98 (18, 11-26) | 23/100 (23, 16-32) | 29/99 (30, 21-39) | 34/98 (35, 26-45) | 44/94 (47, 37-57) | 50/96 (52, 42-62) | 65/95 (68, 59-77) | 62/88 (70, 61-79) |
| | 35-59 | 33/110 (30, 22-39) | 45/110 (41, 32-50) | 52/117 (45, 36-54) | 61/112 (55, 45-64) | 65/111 (59, 49-67) | 70/111 (63, 54-72) | 70/107 (65, 56-74) | 82/104 (79, 70-86) | 82/104 (79, 70-86) |
| | ≥60 | 3/57 (7, 2-14) | 6/55 (12, 5-22) | 7/56 (14, 6-23) | 16/55 (30, 19-42) | 18/53 (35, 23-48) | 23/53 (44, 31-57) | 32/52 (61, 48-74) | 33/51 (64, 51-77) | 30/47 (64, 50-76) |
| | All | 73/505 (14, 12-18) | 109/504 (22, 18-25) | 143/529 (27, 23-31) | 181/510 (36, 31-40) | 209/510 (41, 37-45) | 247/505 (49, 45-53) | 278/499 (56, 51-60) | 339/493 (69, 65-73) | 335/479 (70, 66-74) |

*Adjusted for assay sensitivity and specificity. Blood draw (BD) 1, July 20–September 17, 2020; BD2, September 21–October 10, 2020; BD3, November 23–December 12, 2020; BD4, January 25–February 20, 2021; and BD5, March 22–April 11, 2021; BD6, May 20 – June 9, 2021; BD7, July 19 – August 5, 2021; BD8, September 13 – 25, 2021; BD9, November 15 – 27, 2021.

**Appendix Table 2**. Number and percentage of PHIRST-C participants with known vaccination status (N) vaccinated for SARS-CoV-2 by BD9 (November 15 – 27, 2021) in the rural and urban community, South Africa*

| Age group, y | Rural, No. (%) | | | | Urban, No. (%) | | | |
|---|---|---|---|---|---|---|---|---|
| | N | Not vaccinated | Partially vaccinated | Fully vaccinated | N | Not vaccinated | Partially vaccinated | Fully vaccinated |
| <5 | 93 | 93 (100) | 0 (0) | 0 (0) | 49 | 49 (100) | 0 (0) | 0 (0) |
| 5-12 | 205 | 205 (100) | 0 (0) | 0 (0) | 125 | 125 (100) | 0 (0) | 0 (0) |
| 13-18 | 83 | 82 (99) | 1 (1) | 0 (0) | 78 | 72 (92) | 3 (4) | 3 (4) |
| 19-34 | 101 | 91 (90) | 2 (2) | 8 (8) | 96 | 72 (75) | 8 (8) | 16 (17) |
| 35-59 | 85 | 51 (60) | 9 (11) | 25 (29) | 111 | 59 (53) | 5 (5) | 47 (42) |
| ≥60 | 42 | 16 (38) | 10 (24) | 16 (38) | 53 | 16 (30) | 6 (11) | 31 (58) |
| All | 609 | 538 (88) | 22 (4) | 49 (8) | 512 | 393 (77) | 22 (4) | 97 (19) |

*Partially vaccinated: one dose of Pfizer-BioNTech; Fully vaccinated: one dose of Johnson & Johnson or two doses of Pfizer-BioNTech; at time of blood collection

**Appendix Table 3**. Number and percentage of PHIRST-C participants with either serologic evidence of previous infection (at any draw 20 July, 2020 - November 27, 2021) or fully vaccinated by BD9 (November 15 – 27, 2021) or both in the rural and urban community, South Africa*

| Age group, y | n/N (%) | |
|---|---|---|
| | Rural | Urban |
| <5 | 45/95 (47) | 29/55 (53) |
| 5-12 | 130/208 (63) | 82/132 (62) |
| 13-18 | 62/85 (73) | 68/82 (83) |
| 19-34 | 59/107 (55) | 79/119 (66) |
| 35-59 | 66/90 (73) | 106/122 (87) |
| ≥60 | 27/46 (59) | 47/58 (81) |
| All | 389/631 (62) | 411/568 (72) |

*Fully vaccinated: one dose of Johnson & Johnson or two doses of Pfizer-BioNTech; at time of blood collection

**Appendix Table 4.** The PHIRST-C Group

| Name | Affiliation |
|---|---|
| Amelia Buys | Centre for Respiratory Diseases and Meningitis, National Institute for Communicable Diseases of the National Health Laboratory Service, Johannesburg, South Africa. |
| Anne von Gottberg | Centre for Respiratory Diseases and Meningitis, National Institute for Communicable Diseases of the National Health Laboratory Service, Johannesburg, South Africa. School of Pathology, Faculty of Health Sciences, University of the Witwatersrand, Johannesburg, South Africa. |
| Cheryl Cohen | Centre for Respiratory Diseases and Meningitis, National Institute for Communicable Diseases of the National Health Laboratory Service, Johannesburg, South Africa. School of Public Health, Faculty of Health Sciences, University of the Witwatersrand, Johannesburg, South Africa. |
| F. Xavier Gómez-Olivé | MRC/Wits Rural Public Health and Health Transitions Research Unit (Agincourt), Faculty of Health Sciences, School of Public Health, University of the Witwatersrand, Johannesburg, South Africa. |
| Floidy Wafawanaka | MRC/Wits Rural Public Health and Health Transitions Research Unit (Agincourt), Faculty of Health Sciences, School of Public Health, University of the Witwatersrand, Johannesburg, South Africa. |
| Jackie Kleynhans | Centre for Respiratory Diseases and Meningitis, National Institute for Communicable Diseases of the National Health Laboratory Service, Johannesburg, South Africa. School of Public Health, Faculty of Health Sciences, University of the Witwatersrand, Johannesburg, South Africa. |
| Jacques du Toit | MRC/Wits Rural Public Health and Health Transitions Research Unit (Agincourt), Faculty of Health Sciences, School of Public Health, University of the Witwatersrand, Johannesburg, South Africa. |
| Jinal N. Bhiman | Centre for Respiratory Diseases and Meningitis, National Institute for Communicable Diseases of the National Health Laboratory Service, Johannesburg, South Africa. School of Pathology, Faculty of Health Sciences, University of the Witwatersrand, Johannesburg, South Africa. |
| Jocelyn Moyes | Centre for Respiratory Diseases and Meningitis, National Institute for Communicable Diseases of the National Health Laboratory Service, Johannesburg, South Africa. School of Public Health, Faculty of Health Sciences, University of the Witwatersrand, Johannesburg, South Africa. |
| Kathleen Kahn | MRC/Wits Rural Public Health and Health Transitions Research Unit (Agincourt), Faculty of Health Sciences, School of Public Health, University of the Witwatersrand, Johannesburg, South Africa. |
| Kgaugelo Patricia Kgasago | Perinatal HIV Research Unit (PHRU), University of the Witwatersrand, Johannesburg, South Africa. |
| Limakatso Lebina | Perinatal HIV Research Unit (PHRU), University of the Witwatersrand, Johannesburg, South Africa. Africa Health Research Institute, Durban, South Africa. Africa Health Research Institute, KwaZulu-Natal, South Africa. |
| Linda de Gouveia | Centre for Respiratory Diseases and Meningitis, National Institute for Communicable Diseases of the National Health Laboratory Service, Johannesburg, South Africa. |
| Maimuna Carrim | Centre for Respiratory Diseases and Meningitis, National Institute for Communicable Diseases of the National Health Laboratory Service, Johannesburg, South Africa. School of Pathology, Faculty of Health Sciences, University of the Witwatersrand, Johannesburg, South Africa. |
| Meredith L. McMorrow | Influenza Division, Centers for Disease Control and Prevention, Atlanta, Georgia, United States of America. Influenza Program, Centers for Disease Control and Prevention, Pretoria, South Africa. |
| Mignon du Plessis | Centre for Respiratory Diseases and Meningitis, National Institute for Communicable Diseases of the National Health Laboratory Service, Johannesburg, South Africa. School of Pathology, Faculty of Health Sciences, University of the Witwatersrand, Johannesburg, South Africa. |
| Neil A. Martinson | Perinatal HIV Research Unit (PHRU), University of the Witwatersrand, Johannesburg, South Africa. Johns Hopkins University Center for TB Research, Baltimore, Maryland, United States of America. |

| | |
|---|---|
| Nicole Wolter | Centre for Respiratory Diseases and Meningitis, National Institute for Communicable Diseases of the National Health Laboratory Service, Johannesburg, South Africa. School of Pathology, Faculty of Health Sciences, University of the Witwatersrand, Johannesburg, South Africa. |
| Retshidisitswe Kotane | Centre for Respiratory Diseases and Meningitis, National Institute for Communicable Diseases of the National Health Laboratory Service, Johannesburg, South Africa. School of Pathology, Faculty of Health Sciences, University of the Witwatersrand, Johannesburg, South Africa. |
| Stefano Tempia | Influenza Division, Centers for Disease Control and Prevention, Atlanta, Georgia, United States of America. Influenza Program, Centers for Disease Control and Prevention, Pretoria, South Africa. School of Public Health, Faculty of Health Sciences, University of the Witwatersrand, Johannesburg, South Africa. MassGenics, Atlanta, Georgia, United States of America. |
| Stephen Tollman | MRC/Wits Rural Public Health and Health Transitions Research Unit (Agincourt), School of Public Health, Faculty of Health Science, University of the Witwatersrand. |
| Tumelo Moloantoa | Perinatal HIV Research Unit (PHRU), University of the Witwatersrand, Johannesburg, South Africa. |

**Appendix Figure 1.** Timing of blood collection and district SARS-CoV-2 weekly incidence[1] in a) rural community and b) urban community, March 2020 – November 2021, South Africa.

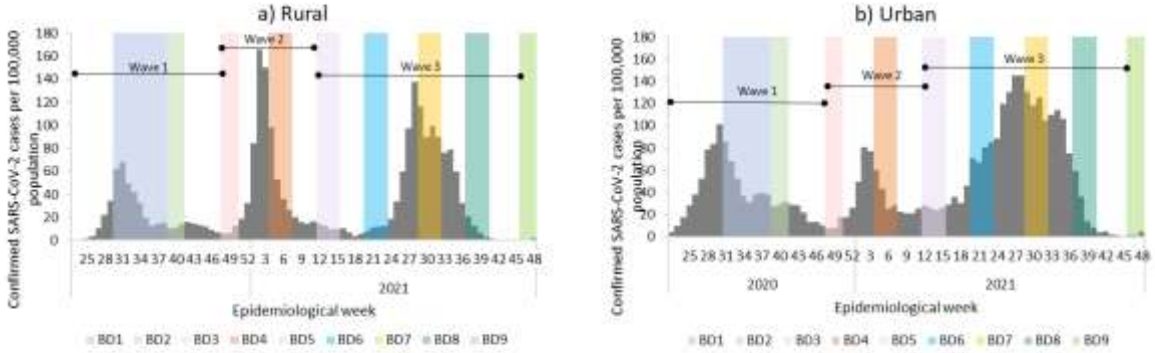


Baseline (BD1) July 20 – September 17, 2020; second draw (BD2) September 21 – October 10, 2020; third draw (BD3) November 23 – December 12, 2020; fourth draw (BD4) January, 25 – February 20, 2021; fifth draw (BD5) March 22 – April 11, 2021; sixth draw (BD6) May 20 – June 9, 2021; seventh draw (BD7) July 19 – August 5, 2021; eighth draw (BD8) September 13 – 25, 2021; ninth draw (BD9) November 15 – 27, 2021. Vertical lines represent 95% credible interval. Seroprevalence estimates adjusted for sensitivity and specificity of assay. Wave 1 (March 1 – November 21, 2020), 2 (November 22, 2020 – March 21, 2021) and 3 (March 22 – November 14, 2021) lines indicate period used for analysis.

[1]Laboratory-confirmed SARS-CoV-2 infections (reverse transcription polymerase chain reaction and antigen tests) reported to the Notifiable Medical Conditions Surveillance System (NMCSS).

**Appendix Figure 2.** South African age-standardized SARS-CoV-2 infection, diagnosis, hospitalization and deaths per 100,000 population during wave 3 in the a) rural and b) urban community, March – November, 2021, South Africa

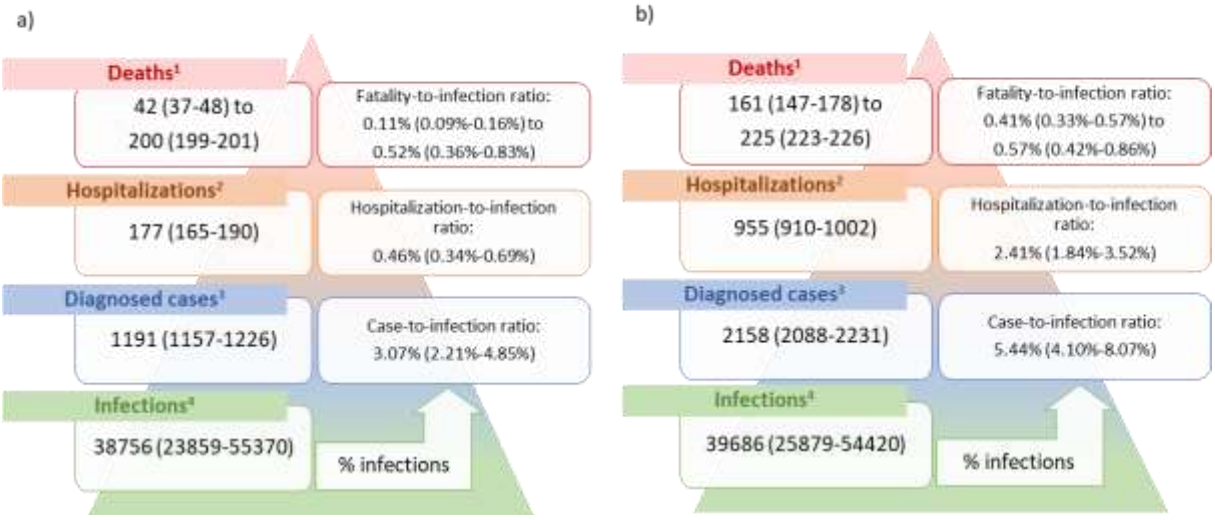


Standardized to South Africa mid-year population estimate for 2021. Wave 3: March 22 – November 14, 2021. [1]Minimum estimate: in-hospitalization deaths in districts based on COVID-19 Sentinel Hospital Surveillance report (DATCOV), maximum estimate: provincial excess deaths reported (South African Medical Research Council). [2]Hospitalizations in the district based on COVID-19 Sentinel Hospital Surveillance (DATCOV). [3]DiagnosedSARS-CoV-2 cases reported from districts from national SARS-CoV-2 surveillance (NMCSS). [1]Based on becoming seropositive or 2-fold increase in cut-off index (COI) between draw 5 and 9. [1]Values in bracket refer to 95% credible interval for infections and 95% confidence interval for all other estimates.

**Appendix Figure 3**. SARS-CoV-2 a) case-to-infection (CIR), hospitalization-to-infection (HIR) and b) in-hospital (IH) fatality-to-infection (FIR) and excess (ED) fatality-to-infection (FIR) ratios in a rural and urban community during the first, second and third wave of infections, March 2020 - November 2021, South Africa.

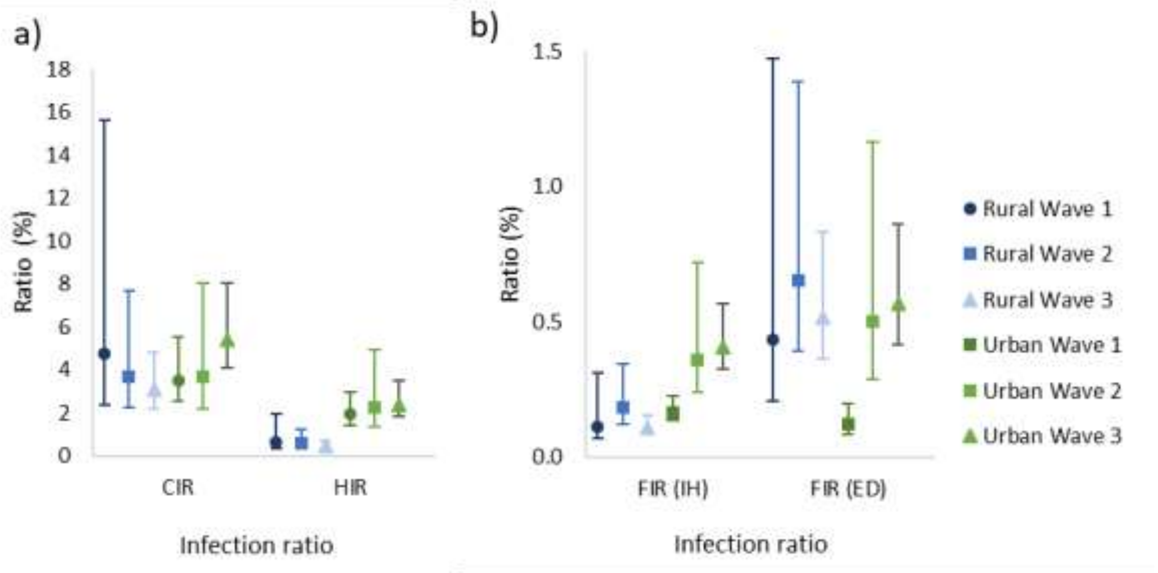
